# Radiomic-Based Approaches in the Multi-metastatic Setting: A Quantitative Review

**DOI:** 10.1101/2024.07.04.24309964

**Authors:** Caryn Geady, Hemangini Patel, Jacob Peoples, Amber Simpson, Benjamin Haibe-Kains

## Abstract

**Background:** Radiomics traditionally focuses on analyzing a single lesion within a patient to extract tumor characteristics, yet this process may overlook inter-lesion heterogeneity, particularly in the multi-metastatic setting. There is currently no established method for combining radiomic features in such settings, leading to diverse approaches with varying strengths and limitations. Our quantitative review aims to illuminate these methodologies, assess their replicability, and guide future research toward establishing best practices, offering insights into the challenges of multi-lesion radiomic analysis across diverse datasets.

**Methods:** We conducted a comprehensive literature search to identify methods for integrating data from multiple lesions in radiomic analyses. We replicated these methods using either the author’s code or by reconstructing them based on the information provided in the papers. Subsequently, we applied these identified methods to three distinct datasets, each depicting a different metastatic scenario.

**Results:** We compared ten mathematical methods for combining radiomic features across three distinct datasets, encompassing a total of 16,850 lesions in 3,930 patients. Performance of these methods was evaluated using the Cox proportional hazards model and benchmarked against univariable analysis of total tumor volume. We observed variable performance in methods across datasets. However, no single method consistently outperformed others across all datasets. Notably, while some methods surpassed total tumor volume analysis in certain datasets, others did not. Averaging methods showed higher median performance in patients with colorectal liver metastases, and in soft tissue sarcoma, concatenation of radiomic features from different lesions exhibited the highest median performance among tested methods.

**Conclusions:** Radiomic features can be effectively selected or combined to estimate patient-level outcomes in multi-metastatic patients, though the approach varies by metastatic setting. Our study fills a critical gap in radiomics research by examining the challenges of radiomic-based analysis in this setting. Through a comprehensive review and rigorous testing of different methods across diverse datasets representing unique metastatic scenarios, we provide valuable insights into effective radiomic analysis strategies.

## Background

Radiological imaging and its quantitative analysis, known as radiomics, aim to transform medical images into mineable data to enhance decision-making in diagnosis, prognosis, and treatment planning. [1]. Traditionally, radiomics has focused on the analysis of a single lesion within a patient, aiming to extract relevant features and understand the tumor’s characteristics [2]. While this practical approach minimizes computational complexity and utilizes simpler mathematical models, relying on a single lesion’s analysis may not provide a complete picture of the disease’s intra- and inter-lesion heterogeneity, particularly in patients with multiple lesions [3].

Currently, there is no established methodology for the optimal combination of radiomic features for patients with multiple lesions [3], [4], which has led to a diverse array of approaches, each offering distinct advantages and trade-offs. While it is common to have multiple approaches in various fields, specific challenges in radiomics warrant a more comprehensive examination. Many studies lack transparency and reproducibility, with methods and code not publicly available, hindering scientific progress [5]. Additionally, there is no comprehensive comparative study evaluating different methods for combining information from multiple lesions in different metastatic scenarios. With advancements in AI and segmentation [6], integrating and comparing multiple lesions has not only become feasible, but also an important problem to solve within the context of precision oncology.

In this study, we quantitatively reviewed the state-of-the-art approaches for multi-lesions radiomic analysis, highlighting their strengths and weaknesses, to guide future research toward establishing best practices in this domain. Serving as a case study in replicability [7], we systematically explore literature methods and implement them across diverse datasets. Our goal is to not only assess the replicability of these methods but also highlight their applicability and effectiveness in addressing the challenges of radiomic analysis in patients with multiple lesions across multiple domains.

## Methods

The design of the study is represented in Figure 1.

**Figure 1:**
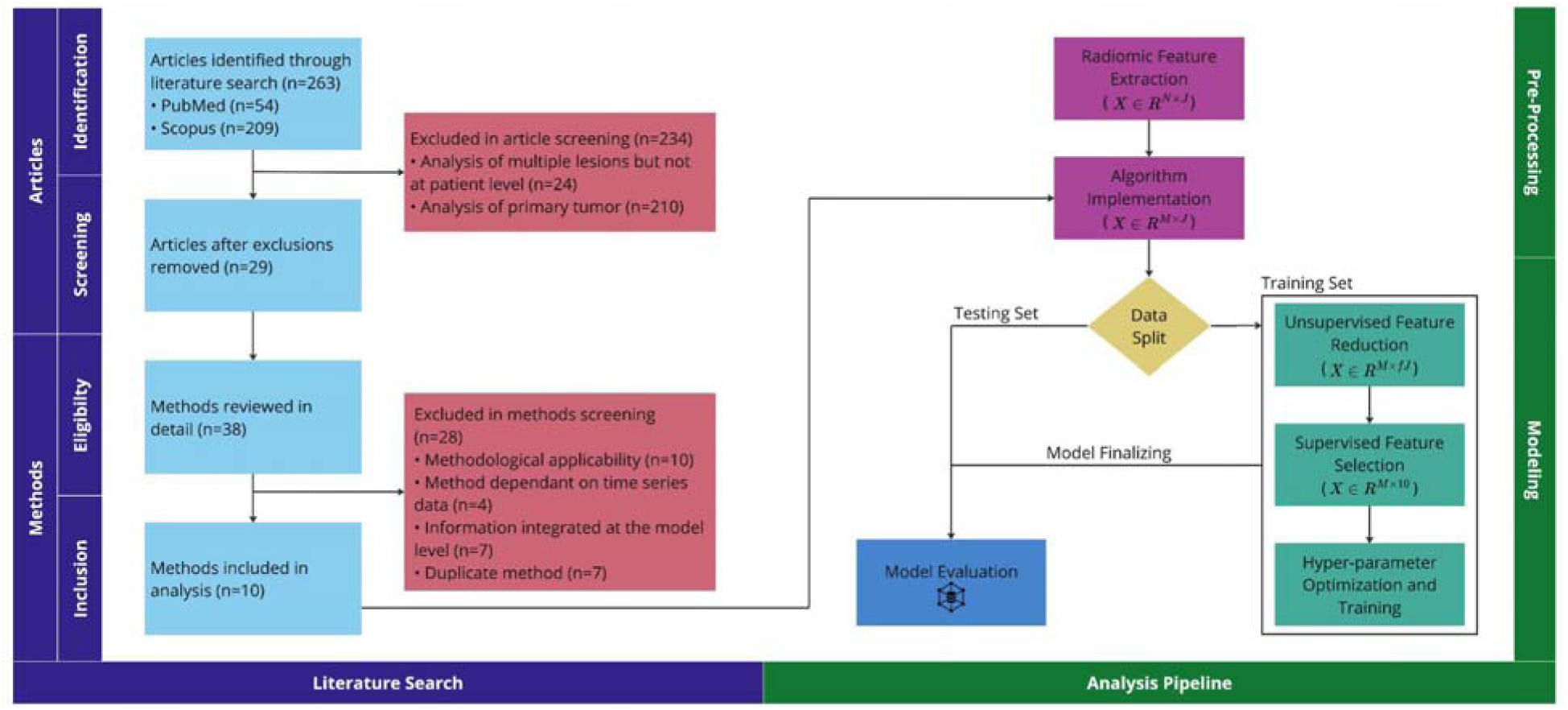
Our study encompasses both a literature search and analysis pipeline. Within the literature search section, inquiries are categorized into article-specific and method-specific, accommodating papers detailing multiple radiomic-based analysis methods in the context of multiple ROIs. The analysis pipeline section is divided into pre-processing and modeling components. Radiomics features, initially forming an N × J lesion-level matrix (where N is the number of lesions and J is the number of radiomic features), are transformed post-method implementation into an M × J patient-level matrix (where M represents the number of patients). Following unsupervised feature reduction, the resulting matrix contains fJ radiomic features, with f representing a fraction within the range [0,1]. Figure was generated using Miro.

### Literature Search

We employed a dual-database methodology, utilizing Scopus and PubMed, to execute a comprehensive and systematic review. Our search strategy involved the incorporation of specific keywords such as radiomics, metastasis, intertumor (lesion), interlesion heterogeneity, feature aggregation, response, and survival, aiming to retrieve a diverse range of scholarly articles. We specifically focused on full-length articles published in English. Detailed queries can be found in the Supplemental section.

#### Selection Criteria

The selection criteria for inclusion focused on the key aspects of patient-level analysis and methodological clarity. Methods were excluded from the review if they met one or more of the exclusion criteria in **Table 1** or were a duplicate method.

**Table 1:** Overview of the methodological exclusion criteria utilized for our quantitative review.

| Exclusion Criterion | Description |
| --- | --- |
| Analysis of multiple lesions but not at patient-level | Multiple lesions were analyzed separately towards tumor-specific predictions; data was not combined/selected/aggregated at the patient level |
| Analysis of primary tumor | Analysis of the primary tumor for prediction of distant or local recurrence or some other patient-level outcome |
| Methodological applicability | Detailed descriptions of employed methods lacking or insufficient and/or code unavailable, presenting significant challenges in method replication and/or proprietary data not available |
| Method dependent on time series data | Measures of treatment response (e.g., change in lesion size at a secondary time point) incorporated into method |
| Information integrated at the model level | Transformations or interactions within a model architecture as opposed to transformations to the raw or pre-processed data used as input to the model |

#### Data Collection

For this pan-cancer review we leveraged three different datasets, each presenting a unique context for conducting radiomic-based analyses among patients with multiple lesions (**Table 2**). The first dataset encompasses primary tumors accompanied by invaded lymph nodes, offering insights into radiomic analysis within the context of localized spread. The second dataset focuses on metastases confined to a single organ, providing an opportunity to explore radiomic patterns specific to localized metastatic disease. In contrast, the third dataset encompasses patients with metastases distributed ubiquitously across the body, allowing for a comprehensive examination of radiomic features in a setting of widespread metastatic involvement. These varied settings enable a multifaceted investigation into radiomic characteristics across different disease extents, contributing to a more comprehensive understanding of radiomic signatures in diverse cancer scenarios.

**Table 2:**
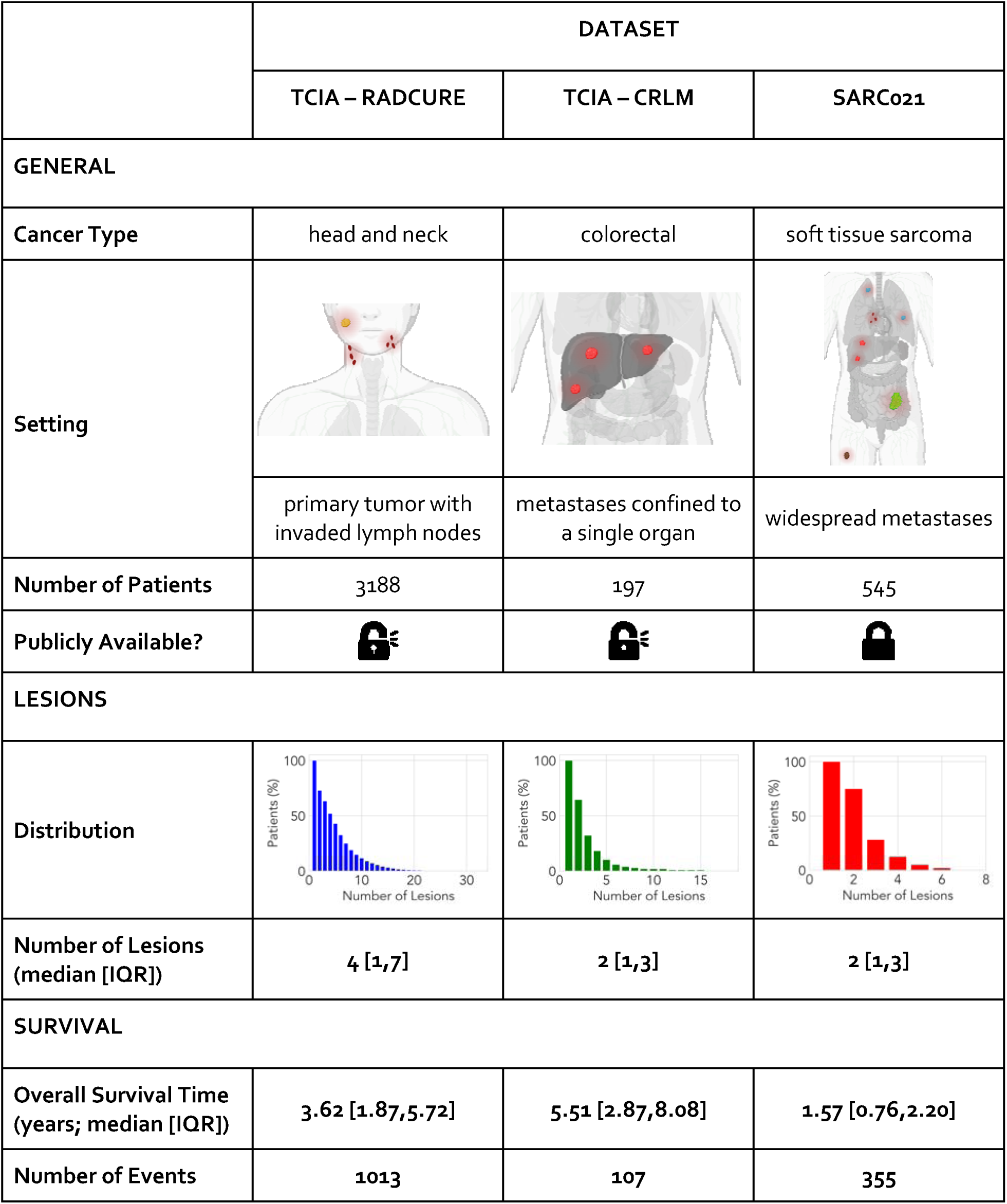
Overview of the datasets utilized in this pan-cancer study. Each dataset represents a uniqu metastatic scenario; two of the datasets are publicly available and one is a private dataset. Statistics surrounding the number of lesions per patient and survival are expressed in terms of the median and th e interquartile range (IQR).

|  | DATASET |  |  |
| --- | --- | --- | --- |
|  | TCIA – RADCURE | TCIA – CRLM | SARCo21 |
| GENERAL |  |  |  |
| Cancer Type | head and neck | colorectal | soft tissue sarcoma |
| Setting |  |  |  |
|  | primary tumor with invaded lymph nodes | metastases confined to a single organ | widespread metastases |
| Number of Patients | 3188 | 197 | 545 |
| Publicly Available? |  |  |  |
| LESIONS |  |  |  |
| Distribution |  |  |  |
| Number of Lesions (median [IQR]) | 4 [1,7] | 2 [1,3] | 2 [1,3] |
| SURVIVAL |  |  |  |
| Overall Survival Time (years; median [IQR]) | 3.62 [1.87,5.72] | 5.51 [2.87,8.08] | 1.57 [0.76,2.20] |
| Number of Events | 1013 | 107 | 355 |

##### TCIA - RADCURE

The Cancer Imaging Archive (TCIA) - RADCURE dataset encompasses data from 3188 patients, comprising computed tomography (CT) images paired with contours delineating primary tumors and invaded lymph nodes. Among the patients analyzed, oropharyngeal cancer accounts for 44% of the population, while larynx, nasopharynx, and hypopharynx cancers constitute 27%, 11%, and 5%, respectively. Additionally, the publicly-available dataset features clinical data linked to each patient, encompassing demographic details, clinical information and survival. Detailed information can be found in [8].

##### TCIA - CRLM

The TCIA - Colorectal-Liver-Metastases (CRLM) dataset comprises CT images and a comprehensive clinical record of 197 patients with colorectal liver tumors. Each patient within this dataset underwent major liver resection as part of their treatment regimen. The CT scan images acquired preoperatively, while the clinical data confines patients’ demographic, pathologic and survival data for each patient. Detailed descriptions can be found in [9].

##### SARC021

SARC021 is a private dataset obtained from the Sarcoma Alliance for Research through Collaboration (SARC). The dataset consists of 545 soft tissue sarcoma patients who were enrolled in a phase III clinical trial (TH-CR-406/SARC021, NCT01440088). Patients presented with either locally advanced, unresectable or metastatic soft tissue sarcoma. Among the patients, leiomyosarcoma accounts for 37% of the population, while liposarcoma, undifferentiated pleomorphic sarcoma and other sarcomas constitute 17%, 12%, and 34%, respectively. Data was collected retrospectively and in accordance with our institutional REB (#20-5707). Collected data includes pre-treatment CT, patient characteristics and survival. Detailed descriptions, including trial protocol and results can be found in [10].

### Algorithm Implementation

Our study utilized 10 different techniques for radiomic analysis of multiple lesions, which can be broadly classified into three categories: lesion selection methods, methods that incorporate information from select lesions, and methods that incorporate information from all lesions (**Table 3)**.

**Table 3:**
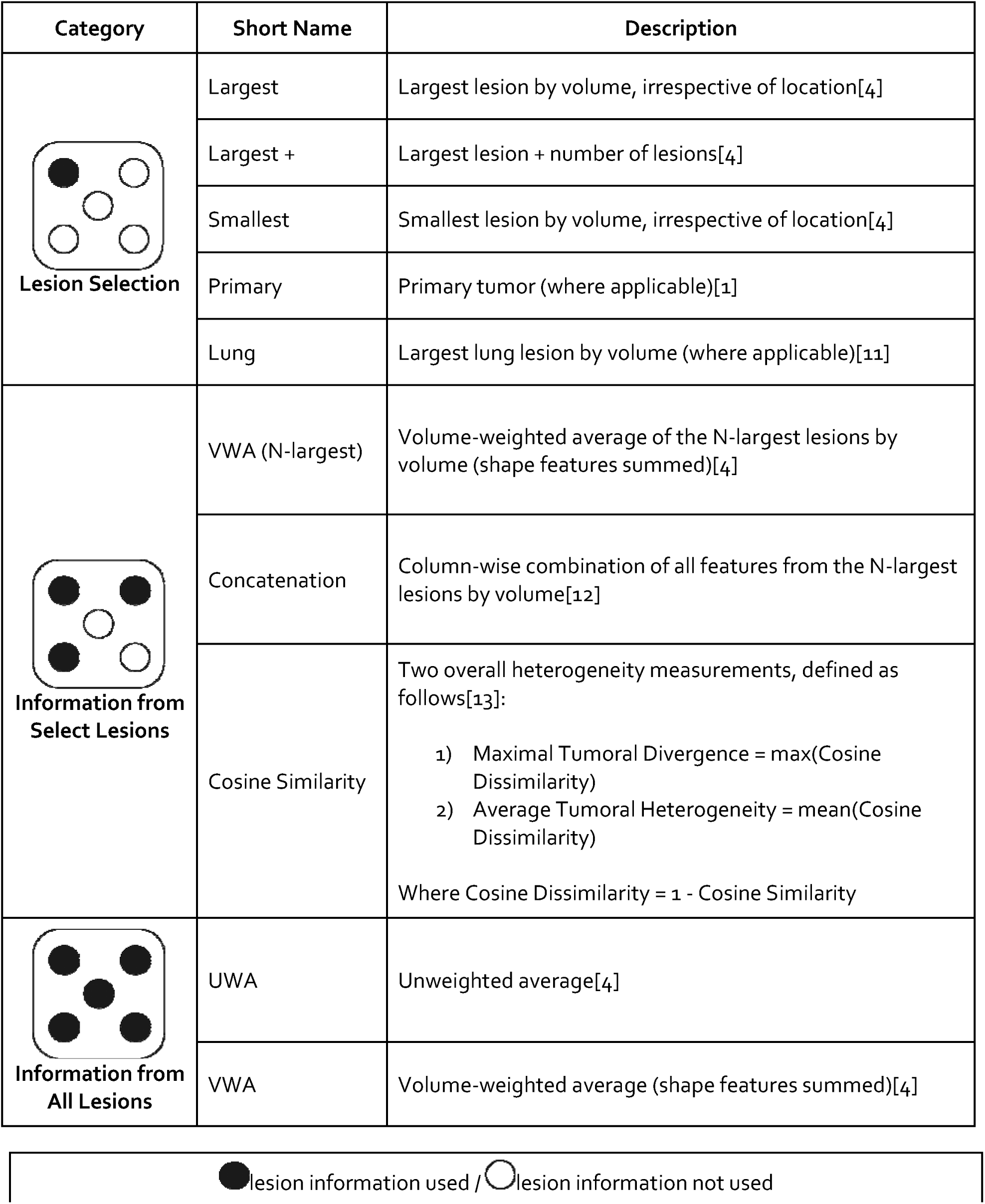
Descriptions of the methods implemented for our analysis.

| Category | Short Name | Description |
| --- | --- | --- |
| **Lesion Selection** | Largest | Largest lesion by volume, irrespective of location[4] |
|  | Largest + | Largest lesion + number of lesions[4] |
|  | Smallest | Smallest lesion by volume, irrespective of location[4] |
|  | Primary | Primary tumor (where applicable)[1] |
|  | Lung | Largest lung lesion by volume (where applicable)[11] |
| **Information from Select Lesions** | VWA (N-largest) | Volume-weighted average of the N-largest lesions by volume (shape features summed)[4] |
|  | Concatenation | Column-wise combination of all features from the N-largest lesions by volume[12] |
|  | Cosine Similarity | Two overall heterogeneity measurements, defined as follows[13]:<br>1) Maximal Tumoral Divergence = max(Cosine Dissimilarity)<br>2) Average Tumoral Heterogeneity = mean(Cosine Dissimilarity)<br>Where Cosine Dissimilarity = 1 - Cosine Similarity |
| **Information from All Lesions** | UWA | Unweighted average[4] |
|  | VWA | Volume-weighted average (shape features summed)[4] |

lesion information used / lesion information not used

#### Lesion Selection Methods

For the proposed lesion selection methods, implementation involved simply selecting a lesion for analysis based on either the lesion label (e.g., primary tumor) or computed lesion volume. For the latter, computation utilized the PyRadiomics library[14], specifically the ‘VoxelVolume’ shape feature. Importantly, lesion label selection was dataset-specific; for instance, the TCIA-RADCURE dataset exclusively contained primary tumors, while SARC021 exclusively contained lung lesions. Consequently, there were variations in sample sizes, with only 93% of patients in the TCIA-RADCURE dataset having primary tumors, and 60% of patients in the SARC021 dataset having at least one lung lesion (**Supplementary Table S1**).

#### Information from Select Lesions

In the proposed combinatorial methods utilizing information from select lesions, we incorporated data from N lesions, where N represents the minimum number of lesions observed in any given patient across all patients.

##### VWA (N-largest)

For the averaging approach, the N-largest lesions were selected. For the patient population where N equals one, this method mirrors the largest lesion approach; however, in groups with two or more lesions, radiomic features from the N-largest lesions are incorporated accordingly. Shape features were summed over selected lesions; all other features constitute a volume-weighted average of selected lesions ((1)).

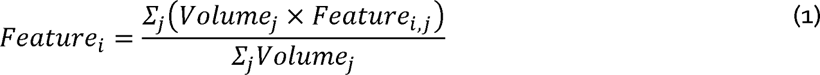

##### Concatenation

For the concatenation approach, the N-largest lesions were selected. Similar to the averaging approach, this method mirrors the largest lesion approach for the patient population where N equals one; however, in groups with two or more lesions, radiomic features from the N-largest lesions are incorporated accordingly. In the case where N equals two, let *A* represent a 1×*n* matrix containing *n* radiomic features for the largest lesion in a given patient. Similarly, let *B* represent a 1×*n* matrix containing n radiomics features for the *second*-largest lesion in the same patient. The matrices *A* and *B* are horizontally concatenated to create *C*, a 1× (2*n*) matrix, containing *n* radiomic features for the largest lesion *and n* radiomics features for the *second*-largest lesion. This can be expanded for the case where N equals three, and so on.

##### Cosine Similarity

For the cosine similarity approach, N-random lesions were utilized to maintain alignment with the analysis in [13]. When N equals one, no pairwise comparisons between lesions are possible. Unsupervised feature reduction was performed prior to calculating the metrics; given the definition of only two metrics (**Table 3**), no supervised feature selection was carried out. Moreover, the cosine similarity method is distinct from other methods as it uses secondary features derived from radiomic features, rather than the radiomic features themselves.

#### Information from All Lesions

For the proposed combinatorial methods utilizing information from all lesions, no modifications or adjustments were made. Lesion volume for the volume-weighted averaging was computed as described above, using the PyRadiomics ‘VoxelVolume’ shape feature.

##### UWA

Shape features were summed over all lesions; all other features constitute an average of all N lesions ((2)).

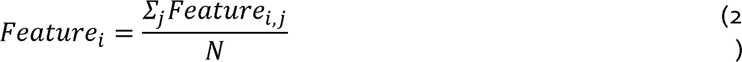

##### VWA

Shape features were summed over all lesions; all other features constitute a volume-weighted average of all *N* lesions (as in (1), but with all *N* lesions).

#### Research Reproducibility

To ensure transparency, reproducibility and reusability of the study outputs, we packaged all the processed data and computer code into a fully-specified software environment to ensure that the results can easily be reproduced. To achieve this, we built a Code Ocean capsule and published it under https://codeocean.com/capsule/7943659/tree.

### Data Analysis

In the context of this study, we have raw data with lesion-level information and a transformed dataset with patient-level information. The lesion-level dataset refers to the raw data that contains detailed information about individual lesions. This dataset is created by extracting radiomic features from all lesions [in all patients] in each dataset, using PyRadiomics [14] (version 3.0.1). Algorithms were implemented on each lesion-level dataset in turn, including lesion selection, combining data from select lesions, and utilizing information from all lesions (**Table 3**). to create patient-level datasets. The patient-level datasets contain the transformed data, that aggregates or summarizes the lesion-level information. Each resultant patient-level dataset was partitioned, stratified by lesion count. Subsequently, unsupervised feature reduction was applied to the training set, where applicable. Initially, a variance filter was employed to eliminate features with less than 10% variance, followed by the removal of radiomic features exhibiting an absolute Spearman rho correlation with lesion volume greater than 0.1[15]. From the resulting reduced feature set, a supervised feature selection technique, specifically mRMR, was utilized to select 10 features[16]. Overall survival analysis employed the Cox proportional hazards model, a widely utilized method for modeling time-to-event data[17]. Optimal hyperparameters were identified by Grid Search[18], employing five-fold cross-validation; the training set was subsequently equipped with these hyperparameters and underwent bootstrapping to ascertain a confidence interval for the concordance index (C-Index)[19]. A total of 100 bootstrap samples were used, with each sample consisting of 50% of the training data. The training hyperparameters were then utilized to train a model on the testing set. Univariable analysis using total lesion volume was used for benchmarking. Key aspects of our study, including the literature search and analysis pipeline, are visualized in **Figure 1**.

#### Subgroup Analysis

A subgroup analysis was conducted to compare model performance across lesion selection and feature aggregation methods by including patients with variable numbers of lesions. Specifically, we examined patients across all lesion counts, as well as exclusive subgroups with two or more lesions, and three or more lesions. This analysis was motivated by the hypothesis that with an increasing number of lesions, the application of certain methods may exhibit performance improvements.

## Results

### Literature Search

Of the 263 articles screened for our study, only 11% met the inclusion criteria for our study (**Figure 1**). Excluded articles fell into one of two categories: (1) those focused on radiomic-based analysis of the primary tumor for predicting local or distant metastasis, and (2) those centered on radiomic-based analysis of a single lesion to predict its response to treatment ((**Figure 1**)). Among the methods reviewed in detail, 26% were included in our study (**Figure 1**). In addition to the exclusion criteria (**Table 1**), redundant methods from one paper that were identical to those from another were eliminated, constituting 7 out of 38 methods.

### Method Implementation

**Figure 2:**
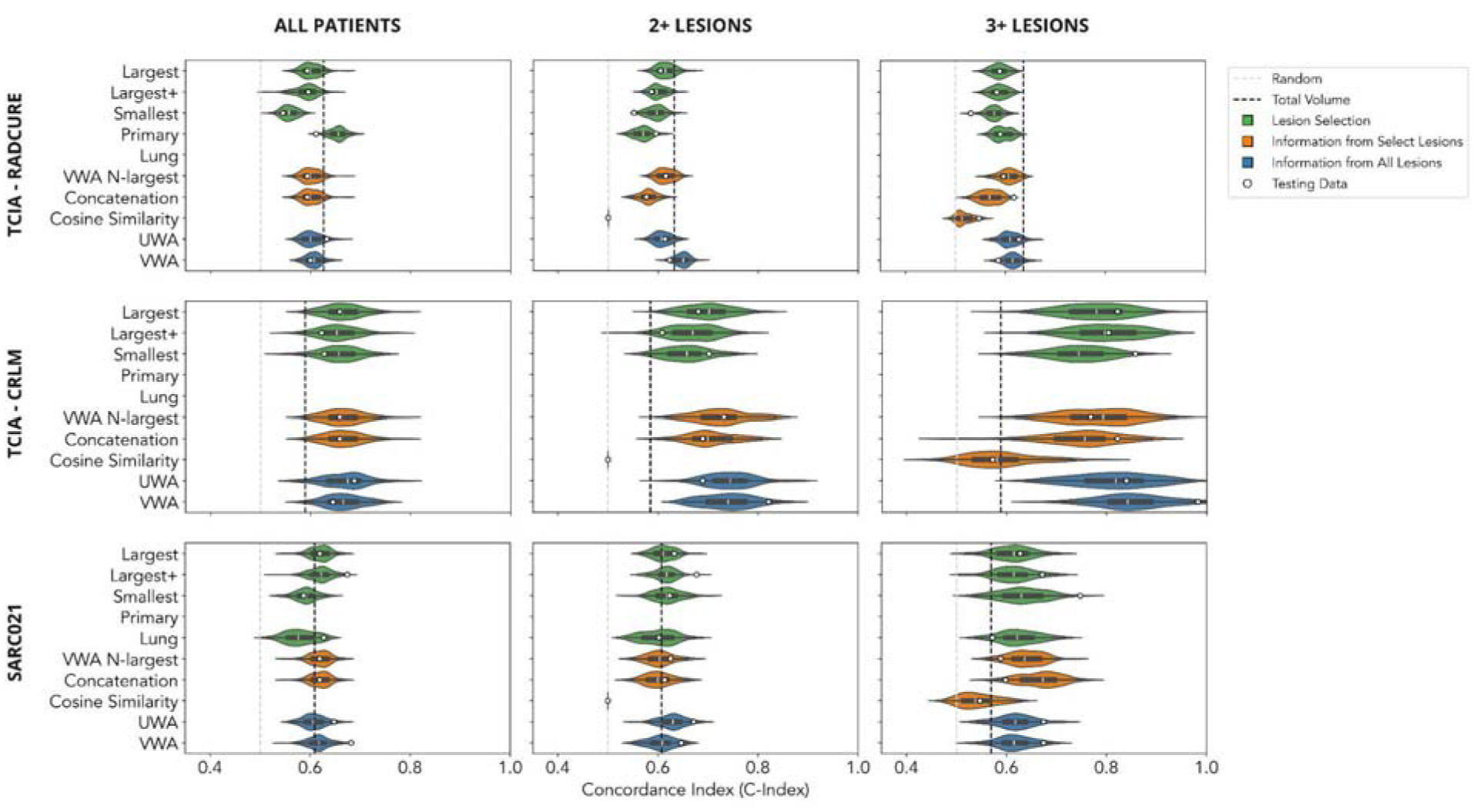
Overall survival analysis, organized by dataset (rows) and subgroup (columns). Implemented methods are color-coded by category. Violin plots depict the performance distribution of bootstrapped samples from the training data, with white circles indicating performance on the testing data. Vertical lines in gray and black represent performance benchmarks from a random model and univariable analysis based on total lesion volume, respectively. Figure was constructed using Miro; numerical results can be found in the Supplemental section.

#### TCIA - RADCURE

When considering all patients, selecting the primary tumor and using 10 features provides the best performance among all implemented methods. For the different subgroups, the primary tumor was the largest lesion in 79%, 73%, and 72% of patients, respectively. This could explain the discrepancy in model performance when comparing radiomic features from the primary tumor versus radiomic features from the largest lesion for all patients. When different subgroups are considered, the performance of the model using radiomic features from the primary tumor worsens. Moreover, no radiomic-based method consistently outperformed univariable analysis of total tumor volume.

#### TCIA - CRLM

In contrast with TCIA - RADCURE, where univariable analysis of total tumor volume consistently outperformed most radiomic methods, in TCIA - CRLM we found that many methods surpassed total tumor volume analysis. This suggests that radiomics-based analysis performs well in this setting. Furthermore, subgroup analysis based on increasing lesion numbers revealed a notable improvement in model performance, underscoring the importance of incorporating information from multiple lesions. When increasing from one lesion to two lesions, the median performance of all models increased (C-Index range [0.001,0.074], with the largest increases observed in the VWA-N-largest and Concatenation methods). In terms of performance distribution, the positive shift was significant for all methods, save for the Smallest Lesion method (two-sample Kolmogorov-Smirnoff test, p-value range [1.42e-19,0.470]). When increasing from two lesions to three lesions, the median performance of all models increased (C-Index range [0.057,0.127], with the largest increases observed in the Largest+ lesion and Concatenation methods). In terms of performance distribution, the positive shift was significant for all methods (two-sample Kolmogorov-Smirnoff test, p-value range [3.054e-31,1.555e-7]). The highest median performance was observed for the VWA method for the three or more lesions subgroup (C-Index 0.842). However, it’s worth noting that as sample sizes decreased, the distribution of model performance widened, indicating potential overfitting. Additionally, averaging methods exhibited higher median values in terms of model performance.

#### SARC021

In SARC021, we observed a mixed pattern of results. Similar to TCIA - RADCURE, no radiomic-based method consistently outperformed univariable analysis of total tumor volume when considering all patients and the subgroup of patients with 2 or more lesions. However, akin to TCIA - CRLM, we noted a marked shift in overall model performance in the subgroup of patients with 3 or more lesions, albeit with less variability. When increasing from two lesions to three lesions, the median performance of all but two models increased (C-Index range [-0.012,0.073], with the largest increases observed in the Concatenation method). In terms of performance distribution, the positive shift was significant for all methods except the Largest, Largest+ and VWA methods (two-sample Kolmogorov-Smirnoff test, p-value range [4.959e-43,0.111]). The highest median performance was observed for the Concatenation method for the three or more lesions subgroup (C-Index 0.673).

## Discussion

Despite the prevalence of multi-metastatic patients, most radiomic studies have traditionally focused on a single lesion. At time of publication, a search on PubMed yielded around 12,000 hits for radiomics, yet only 38 articles met the criteria for this study, highlighting the need for more comprehensive research. Moreover, there is currently no established methodology for effectively combining radiomic features from multiple lesions [3], [4]. To address this gap, we conducted a comprehensive quantitative review of radiomic-based analyses involving multiple lesions. We replicated and applied 10 different methods across three distinct datasets, each representing a unique metastatic scenario. By collating and evaluating various methods from the literature, our study provides valuable insights into what works, and what does not, in different clinical scenarios. This paper serves as a resource for radiomics researchers, offering guidance for implementing similar techniques in their own data and serving as a foundation for further exploration in this field.

Our findings within the setting of localized metastatic spread align with those of [20], where the top-performing model incorporated clinical features and tumor volume; although [20] focused on the primary tumor and used a subset of our dataset, our results complement theirs by offering a broader analysis across multiple lesions. Our study illustrates that no radiomic-based method combining information from multiple lesions consistently outperformed univariable analysis based on total tumor volume. In this setting, our findings suggest that adhering to the conventional approach of analyzing the primary tumor yields optimal results across all patients, albeit with the acknowledgment that total tumor volume, a simpler measure, performs comparably well.

Our findings within the setting of metastases confined to a single organ align with those of a previous study [4]. In both studies, averaging methods demonstrated optimal performance. Although the previous study focused on brain metastases and utilized magnetic resonance imaging, while our study centered on colorectal liver metastases and employed computed tomography, the similarity in the effectiveness of the methods is noteworthy. In the context of colorectal liver metastases, the number of tumors has been identified as a prognostic factor [21], which could potentially explain the observed quantitative shift in model performance when analyzing subgroups with increasing numbers of lesions. Even so, our analysis revealed greater variability in model performance with decreasing sample sizes, indicating potential overfitting and underscoring the need for cautious interpretation of results in smaller datasets.

Given the limited precedent in radiomics for sarcoma, particularly in the context of widespread, multi-metastatic scenarios, our study was presented with unique analytical challenges and opportunities. We incorporated the largest lung lesion method into our analysis, recognizing the lung as a common metastatic site in sarcomas [11]. Across all patients, the largest lesion method showed marginally improved performance compared to univariable analysis of total tumor volume, highlighting incremental gains. However, the most intriguing findings surfaced when examining the subgroup of patients with 3 or more lesions. Here, nearly all methods demonstrated median model performance surpassing that of total tumor volume, with the concatenation method standing out with the highest median performance. These results suggest that in multi-metastatic scenarios, individual lesions may harbor unique prognostic information that could potentially enhance prognostic accuracy beyond traditional approaches relying solely on total tumor volume.

One method of interest is the cosine similarity metrics originally proposed for characterizing inter-tumoral heterogeneity, described in [13], which holds promise from both theoretical and applied perspectives. However, its effectiveness is hindered by limitations in lesion sampling. In the original study, the authors found that sampling 2 or 3 lesions at random recovered 48% and 67% of the average tumoral heterogeneity, respectively, and 27% and 55% of the maximal tumor divergence, respectively. However, the authors emphasized that "more than eight lesions per patient would have been necessary to recover at least 75% of the true heterogeneity captured by the whole lesion distribution analysis" [13]. Unfortunately, such extensive sampling is often not feasible due to limited availability of patients with multiple lesions sampled. Furthermore, the underwhelming performance of this method in our study may also be attributed to the limited number of features used in the model. While only two features were defined for this method, our analysis employed 10 features for all other methods, potentially impacting its comparative performance.

Our study has several potential limitations. From a methodological standpoint, our study relates to the sample size and the approach to feature selection. We aimed to maintain consistency across all datasets by employing a standardized process, including the use of 10 imaging features from each method, with the exception of the cosine method. This decision was made to ensure comparability of results across different datasets. However, the choice of 10 features was influenced by the number of events in the TCIA - CRLM dataset, which had 107 events. While it may have been feasible to use more features for datasets with a higher number of events, we opted to err on the side of caution to limit overfitting or the introduction of bias in this specific dataset. Our study also faced challenges in terms of replicability, largely due to the limited availability of code for the methods included in our analysis. Unfortunately, this reflects a broader issue within research, where adherence to open science standards varies. Despite this obstacle, we endeavored to mitigate replicability concerns by developing modular functions closely aligned with the literature and making them publicly available. This approach not only enhances transparency but also enables future researchers to easily adapt and utilize our methods for their own analyses, thus contributing to the advancement of open science practices in radiomics research.

## Conclusion

Our study fills a critical gap in radiomics research by examining the challenges of analyzing multiple lesions in multi-metastatic patients. Through a comprehensive review and rigorous testing of different methods across diverse datasets representing unique metastatic scenarios, we provide valuable insights into effective radiomic analysis strategies. Moreover, our publicly available code on GitHub and Code Ocean enhances reproducibility and serves as a valuable resource for researchers in the field.

## Data Availability

All data produced are available online at https://codeocean.com/capsule/7943659/tree.

https://www.cancerimagingarchive.net/collection/colorectal-liver-metastases/

https://www.cancerimagingarchive.net/collection/radcure/

## List of Abbreviations

TCIA: The Cancer Imaging Archive
CT: Computed Tomography
CRLM: Colorectal Liver Metastases
SARC: Sarcoma Alliance for Research through Collaboration
mRMR: minimum Redundancy Maximum Relevance
C-Index: Concordance Index

## Declarations

### Ethics approval and consent to participate

The TCIA - RADCURE and TCIA - CRLM datasets are both publicly available; therefore, there was no need to acquire approval or consent. The SARC021 dataset was acquired through Data Use Agreements (DUA) from the Sarcoma Alliance for Research through Collaboration (SARC, https://sarctrials.org/). Consent was waived for our retrospective study through the University Health Network (UHN) Research Ethics Board (REB; #20-5707) as our use of these patients’ data was in accordance with their stated preferences when they consented for the trial.

### Consent for publication

Not applicable.

### Availability of data and materials

The imaging data with accompanying segmentations and clinical metadata for the TCIA - RADCURE (<u>10.7937/J47W-NM11</u>) and TCIA - CRLM (<u>10.7937/QXK2-QG03</u>) datasets are both publicly available. Imaging and clinical data for the SARC021 trial are not publicly available but can be obtained through SARC with a DUA (Contact for details on using data from SARC). The de-identified radiomic features dataset along with the code for data cleaning and analysis that underlie the results reported in the article are publicly available. The computer code is available from https://github.com/bhklab/Quantitative-Review. The full software environment, processed data and computer code are available from https://codeocean.com/capsule/7943659/tree.

### Competing interests

The authors declare that they have no competing interests.

### Funding

Research reported in this publication was supported by the National Cancer Institute of the National Institutes of Health under Award Number P50CA272170. The content is solely the responsibility of the authors and does not necessarily represent the official views of the National Institutes of Health.

### Authors’ contributions

CG was responsible for conceptualization, methodology, formal analysis, investigation, data curation, writing the original draft, and visualization. HSP and JP contributed to investigation and data curation. AS handled data and computational resources, and supervision. BHK contributed to conceptualization, data and computational resources, supervision, and funding acquisition. All authors reviewed the manuscript.

## Acknowledgements

We would like to thank SARC for generously providing their data for use in this study. Additionally, we extend our gratitude to members of the BHKLab, and in particular, Katy Scott, for their valuable input and expertise in developing the visualizations used in this research.

## Authors’ information (optional)

N/A

## Supplemental

### Literature Search (Specific Database Queries)

#### Scopus

⍰ TITLE-ABS (radiomics AND (metastas?s OR intertumor) AND (response OR survival)) AND (LIMIT-TO (SUBJAREA, "MEDI")) AND (LIMIT-TO (DOCTYPE, "ar")) AND (LIMIT-TO (LANGUAGE, "English")) AND (LIMIT-TO (EXACTKEYWORD, "Radiomics"));

#### PubMed

⍰ (radiomics AND inter lesion heterogeneity);
⍰ (radiomics AND metastases AND interlesion);
⍰ (radiomics AND feature aggregation);

### Summary of Sample Sizes (Subgroup Analysis)

**Table S1:**
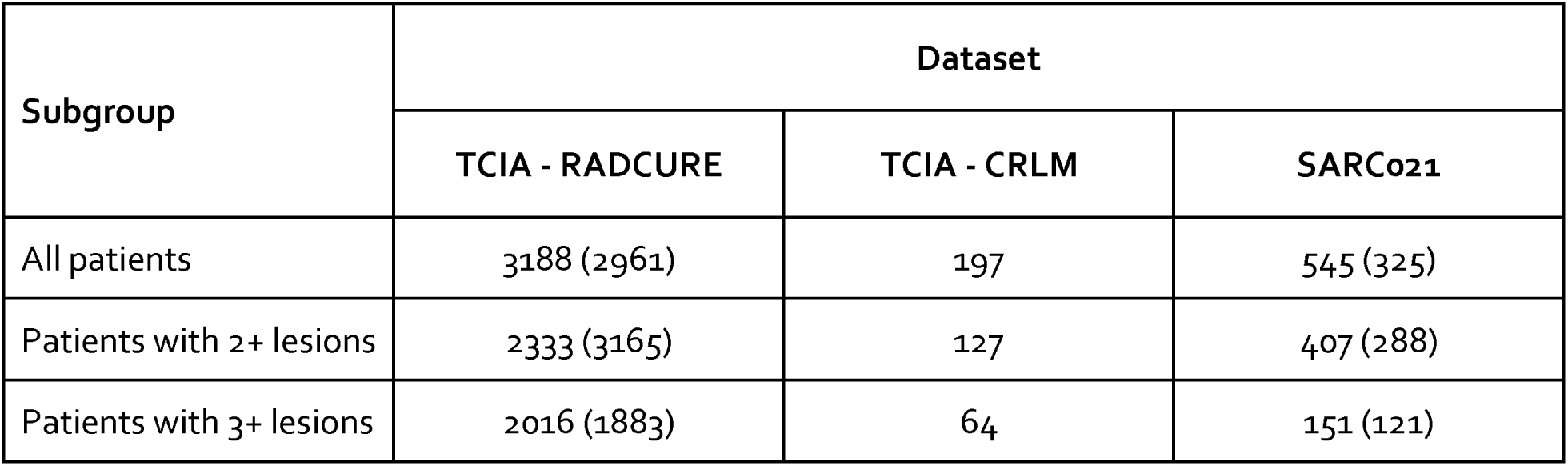
Summary of sample sizes for various subgroup analyses. For the TCIA - RADCURE and SARC021 datasets, numbers in parentheses represent the number of patients with a primary tumor and with a lung metastasis, respectively.

| Subgroup | Dataset |  |  |
| --- | --- | --- | --- |
|  | TCIA - RADCURE | TCIA - CRLM | SARCo21 |
| All patients | 3188 (2961) | 197 | 545 (325) |
| Patients with 2+ lesions | 2333 (3165) | 127 | 407 (288) |
| Patients with 3+ lesions | 2016 (1883) | 64 | 151 (121) |

### Numerical Results

**Table S2:** Comparison of feature aggregation methods for all patients, irrespective of number of tumors, in three distinct metastatic settings. The Cox proportional hazards model was used to fit the data. The training set was bootstrapped to generate confidence intervals (CI) for model performance; the metric used for model performance is the Concordance Index (C-Index).

|  | RADCURE (n=3188) |  | TCIA-CRLM (n=197) |  | SARCo21 (n=545) |  |
| --- | --- | --- | --- | --- | --- | --- |
|  | Train (Median (95% CI))<br>(n=2550) | Test<br>(n=638) | Train (Median (95% CI))<br>(n=157) | Test<br>(n=40) | Train (Median (95% CI))<br>(n=436) | Test<br>(n=109) |
| **Largest Lesion** | 0.599 (0.570-0.634) | 0.593 | 0.664 (0.601-0.739) | 0.658 | 0.622 (0.579-0.659) | 0.619 |
| **Largest Lesion**<br>**+ Number of Mets** | 0.595 (0.549-0.630) | 0.595 | 0.653 (0.583-0.733) | 0.622 | 0.621 (0.577-0.659) | 0.675 |
| **Smallest Lesion** | 0.556 (0.530-0.585) | 0.545 | 0.656 (0.575-0.732) | 0.627 | 0.593 (0.544-0.633) | 0.587 |
| **Primary Tumor\*** | 0.656 (0.625-0.683) | 0.611 | - | - | - | - |
| **Largest Lung Lesion\*** | - | - | - | - | 0.576 (0.523-0.632) | 0.627 |
| **Weighted Average (N-**<br>**largest)** | 0.599 (0.570-0.634) | 0.593 | 0.664 (0.601-0.739) | 0.658 | 0.622 (0.579-0.659) | 0.619 |
| **Concatenation** | 0.599 (0.570-0.634) | 0.593 | 0.664 (0.601-0.739) | 0.658 | 0.622 (0.579-0.659) | 0.619 |
| **Cosine Metrics** | N/A |  |  |  |  |  |
| **Unweighted Average** | 0.600 (0.572-0.645) | 0.632 | 0.675 (0.577-0.752) | 0.687 | 0.605 (0.567-0.653) | 0.648 |
| **Weighted Average** | 0.607 (0.577-0.640) | 0.600 | 0.665 (0.594-0.737) | 0.644 | 0.618 (0.576-0.662) | 0.682 |

**Table S3:** Comparison of feature aggregation methods for patients with two or more tumors, in three distinct metastatic settings. The Cox proportional hazards model was used to fit the data. The training set was bootstrapped to generate confidence intervals (CI) for model performance; the metric used for model performance is the Concordance Index (C-Index).

|  | RADCURE (n=2333) |  | TCIA-CRLM (n=127) |  | SARCo21 (n=407) |  |
| --- | --- | --- | --- | --- | --- | --- |
|  | Train (Median (95% CI))<br>(n=1868) | Test<br>(n=465) | Train (Median (95% CI))<br>(n=102) | Test<br>(n=25) | Train (Median (95% CI))<br>(n=323) | Test<br>(n=84) |
| **Largest Lesion** | 0.614 (0.584-0.649) | 0.604 | 0.702 (0.623-0.785) | 0.681 | 0.611 (0.571-0.649) | 0.633 |
| **Largest Lesion**<br>**+ Number of Mets** | 0.597 (0.572-0.630) | 0.587 | 0.669 (0.583-0.747) | 0.609 | 0.617 (0.573-0.649) | 0.677 |
| **Smallest Lesion** | 0.597 (0.563-0.628) | 0.551 | 0.658 (0.578-0.741) | 0.702 | 0.618 (0.576-0.674) | 0.623 |
| **Primary Tumor\*** | 0.570 (0.536-0.605) | 0.595 | - | - | - | - |
| **Largest Lung Lesion\*** | - | - | - | - | 0.605 (0.539-0.667) | 0.602 |
| **Weighted Average (N-**<br>**largest)** | 0.613 (0.587-0.642) | 0.615 | 0.727 (0.638-0.826) | 0.732 | 0.603 (0.556-0.659) | 0.625 |
| **Concatenation** | 0.579 (0.543-0.612) | 0.576 | 0.699 (0.613-0.797) | 0.689 | 0.600 (0.550-0.658) | 0.614 |
| **Cosine Metrics** | 0.500 (0.500-0.500) | 0.500 | 0.500 (0.500-0.531) | 0.500 | 0.500 (0.500-0.500) | 0.500 |
| **Unweighted Average** | 0.607 (0.578-0.643) | 0.614 | 0.745 (0.656-0.858) | 0.689 | 0.630 (0.583-0.679) | 0.671 |
| **Weighted Average** | 0.650 (0.619-0.670) | 0.623 | 0.740 (0.653-0.842) | 0.821 | 0.609 (0.556-0.656) | 0.646 |

**Table S4:** Comparison of feature aggregation methods for patients with three or more tumors, in three distinct metastatic settings. The Cox proportional hazards model was used to fit the data. The training set was bootstrapped to generate confidence intervals (CI) for model performance; the metric used for model performance is the Concordance Index (C-Index).

|  | RADCURE (n=2016) |  | TCIA-CRLM (n=64) |  | SARCo21 (n=151) |  |
| --- | --- | --- | --- | --- | --- | --- |
|  | Train (Median (95% CI))<br>(n=1613) | Test<br>(n=403) | Train (Median (95% CI))<br>(n=51) | Test<br>(n=13) | Train (Median (95% CI))<br>(n=121) | Test<br>(n=30) |
| **Largest Lesion** | 0.587 (0.558-0.617) | 0.589 | 0.779 (0.659-0.886) | 0.821 | 0.615 (0.532-0.690) | 0.627 |
| **Largest Lesion**<br>**+ Number of Mets** | 0.587 (0.557-0.617) | 0.582 | 0.797 (0.674-0.912) | 0.804 | 0.615 (0.525-0.690) | 0.672 |
| **Smallest Lesion** | 0.578 (0.544-0.609) | 0.531 | 0.744 (0.625-0.849) | 0.857 | 0.630 (0.549-0.719) | 0.748 |
| **Primary Tumor\*** | 0.594 (0.566-0.626) | 0.590 | - | - | - | - |
| **Largest Lung Lesion\*** | - | - | - | - | 0.622 (0.550-0.704) | 0.572 |
| **Weighted Average (N-**<br>**largest)** | 0.606 (0.568-0.637) | 0.596 | 0.793 (0.653-0.909) | 0.768 | 0.636 (0.570-0.693) | 0.588 |
| **Concatenation** | 0.568 (0.530-0.603) | 0.617 | 0.756 (0.553-0.864) | 0.821 | 0.673 (0.586-0.739) | 0.598 |
| **Cosine Metrics** | 0.520 (0.496-0.555) | 0.516 | 0.595 (0.456-0.756) | 0.554 | 0.532 (0.473-0.605) | 0.625 |
| **Unweighted Average** | 0.609 (0.574-0.645) | 0.627 | 0.818 (0.670-0.939) | 0.839 | 0.618 (0.544-0.701) | 0.674 |
| **Weighted Average** | 0.615 (0.581-0.648) | 0.586 | 0.842 (0.714-0.982) | 0.982 | 0.615 (0.545-0.681) | 0.674 |

## Notes

### Competing Interest Statement

The authors have declared no competing interest.

### Author Declarations

Research Ethics Board (REB; #20-5707) of University Health Network (UHN) waived ethical approval for this work.

## References

[1] H. J. W. L. Aerts et al., “Decoding tumour phenotype by noninvasive imaging using a quantitative radiomics approach,” Nat. Commun., vol. 5, p. 4006, Jun. 2014.

[2] J. E. van Timmeren, D. Cester, S. Tanadini-Lang, H. Alkadhi, and B. Baessler, “Radiomics in medical imaging-‘how-to’ guide and critical reflection,” Insights Imaging, vol. 11, no. 1, p. 91, Aug. 2020.

[3] R. Sun, M. Lerousseau, and J. Briend-Diop, “Radiomics to evaluate interlesion heterogeneity and to predict lesion response and patient outcomes using a validated signature of CD8 cells in advanced …,” for ImmunoTherapy of …, 2022, [Online]. Available: https://jitc.bmj.com/content/10/10/e004867.abstract

[4] E. Chang et al., “Comparison of radiomic feature aggregation methods for patients with multiple tumors,” Sci. Rep., vol. 11, no. 1, p. 9758, May 2021.

[5] B. Haibe-Kains et al., “Transparency and reproducibility in artificial intelligence,” Nature, vol. 586, no. 7829. pp. E14–E16, Oct. 2020.

[6] J. Ma and B. Wang, “Towards foundation models of biological image segmentation,” Nat. Methods, vol. 20, no. 7, pp. 953–955, Jul. 2023.

[7] H. E. Plesser, “Reproducibility vs. Replicability: A Brief History of a Confused Terminology,” Front. Neuroinform., vol. 11, p. 76, 2017.

[8] Welch, M. L., Kim, S., Hope, A., Huang, S. H., Lu, Z., Marsilla, J., Kazmierski, M., Rey-McIntyre, K., Patel, T., O’Sullivan, B., Waldron, J., Kwan, J., Su, J., Soltan Ghoraie, L., Chan, H. B., Yip, K., Giuliani, M., Princess Margaret Head And Neck Site Group, Bratman, S., Tadic, T, “Computed tomography images from large head and neck cohort (RADCURE) - the cancer imaging archive (TCIA) public access - cancer imaging archive wiki.” Accessed: Nov. 17, 2023. [Online]. Available: https://wiki.cancerimagingarchive.net/pages/viewpage.action?pageId=70226325

[9] A. L. Simpson et al., “Preoperative CT and survival data for patients undergoing resection of colorectal liver metastases,” Sci Data, vol. 11, no. 1, p. 172, Feb. 2024.

[10] W. D. Tap et al., “Doxorubicin plus evofosfamide versus doxorubicin alone in locally advanced, unresectable or metastatic soft-tissue sarcoma (TH CR-406/SARC021): an international, multicentre, open-label, randomised phase 3 trial,” Lancet Oncol., vol. 18, no. 8, pp. 1089–1103, Aug. 2017.

[11] M. R. Tomaszewski et al., “AI-Radiomics Can Improve Inclusion Criteria and Clinical Trial Performance,” Tomography, vol. 8, no. 1, pp. 341–355, Feb. 2022.

[12] G.-W. Ji et al., “Machine-learning analysis of contrast-enhanced CT radiomics predicts recurrence of hepatocellular carcinoma after resection: A multi-institutional study,” EBioMedicine, vol. 50, pp. 156–165, Dec. 2019.

[13] T. Henry et al., “Investigation of radiomics based intra-patient inter-tumor heterogeneity and the impact of tumor subsampling strategies,” Sci. Rep., vol. 12, no. 1, p. 17244, Oct. 2022.

[14] J. J. M. van Griethuysen et al., “Computational Radiomics System to Decode the Radiographic Phenotype,” Cancer Res., vol. 77, no. 21, pp. e104–e107, Oct. 2017.

[15] A. Traverso et al., “Machine learning helps identifying volume-confounding effects in radiomics,” Phys. Med., vol. 71, pp. 24–30, Mar. 2020.

[16] mrmr: mRMR (minimum-Redundancy-Maximum-Relevance) for automatic feature selection at scale Github. Accessed: Apr. 11, 2024. [Online]. Available: https://github.com/smazzanti/mrmr

[17] J. D. Kalbfleisch and D. E. Schaubel, Fifty Years of the Cox Model. SSRN, 2023.

[18] B. H. Shekar and G. Dagnew, “Grid Search-Based Hyperparameter Tuning and Classification of Microarray Cancer Data,” in 2019 Second International Conference on Advanced Computational and Communication Paradigms (ICACCP), IEEE, Feb. 2019, pp. 1–8.

[19] A. R. Brentnall and J. Cuzick, “Use of the concordance index for predictors of censored survival data,” Stat. Methods Med. Res., vol. 27, no. 8, pp. 2359–2373, Aug. 2018.

[20] M. Kazmierski et al., “Multi-institutional Prognostic Modeling in Head and Neck Cancer: Evaluating Impact and Generalizability of Deep Learning and Radiomics,” Cancer Res Commun, vol. 3, no. 6, pp. 1140–1151, Jun. 2023.

[21] Y. Fong, J. Fortner, R. L. Sun, M. F. Brennan, and L. H. Blumgart, “Clinical score for predicting recurrence after hepatic resection for metastatic colorectal cancer: analysis of 1001 consecutive cases,” Ann. Surg., vol. 230, no. 3, pp. 309–18; discussion 318–21, Sep. 1999.

[22] L. Cavinato et al., “Radiomics-based inter-lesion relation network to describe [18F]FMCH PET/CT imaging phenotypes in prostate cancer,” Cancers, vol. 15, no. 3, p. 823, Jan. 2023.

[23] H. A. Vargas et al., “A novel representation of inter-site tumour heterogeneity from pre-treatment computed tomography textures classifies ovarian cancers by clinical outcome,” Eur. Radiol., vol. 27, no. 9, pp. 3991–4001, Sep. 2017.

